# Using consideRATE to Evaluate Patient Experience in a Cancer Center: Psychometric and Healthcare Assessments

**DOI:** 10.1101/2025.01.24.25321099

**Authors:** Joseph P. Nano, Gabrielle Stevens, Glyn Elwyn, Leeza Petrov, Saahithya Gowrishankar, Shaday Robitaille, Aricca D. Van Citters, Eugene C. Nelson, Garrett T. Wasp, Kathryn B. Kirkland, Meredith A. MacMartin, Catherine H. Saunders

## Abstract

**Background:** **conside**RATE is a patient- and care-partner-reported measure of care experience during serious illness. We used **conside**RATE with patients and care partners at the Dartmouth Cancer Center to assess patient experience, evaluate psychometric properties, and explore scoring approaches.

**Methods:** Patients and care partners aged 18+ and English proficient participated in a cross-sectional survey. Participants completed **conside**RATE (8 items), CANHELP-Lite (21 items), and demographic questions. Reliability was assessed using Cronbach’s α, and validity was evaluated with Pearson’s correlations. Continuous and top-box scoring approaches were used. Psychometric properties were analyzed for patients, care partners, and subgroups with lower educational attainment or income.

**Results:** 244 participants (114 patients,128 care-partners, 2 unspecified) completed the survey. **conside**RATE has internal reliability (α = 0.86); the correlation (r) between **conside**RATE continuous scoring and CANHELP Lite scores for all participants was 0.5; p<0.001, for patients 0.5; p<0.001, for care-partners 0.5; p<0.001, for patients (n=71) with lower educational attainment 0.5, p<0.001, and for patients (n=50) with lower income 0.7, p<0.001. We also found correlations between **conside**RATE top-box scoring and CANHELP Lite scores for all participants (rpb=0.4, p<0.001), with stronger associations among the patient (rpb=0.5, p<0.001) and lower income (rpb=0.4, p=0.005) subgroups. We found discriminant validity between **conside**RATE and Single-item Health Literacy (SILs) measures for continuous scoring (r = -0.05 to 0.09, p>0.05) and top-box scoring (r = -0.02 to 0.09, p>0.05). We found no significant difference in overall experience between patients with breast and blood cancer.

**Conclusion:** We demonstrated in this sample of patients attending a cancer center that **conside**RATE has good internal reliability and is well correlated with CANHELP-Lite.

## Background

Patients with cancer have a constellation of needs that are often incompletely addressed.^1,2^ Measurement of patient-reported outcomes in oncology is a valuable tool for quality assessment and improvement that can be deployed at both the individual and population levels.^3^ The National Comprehensive Cancer Network (NCCN) Quality and Outcomes Committee developed a list of priority concepts, quality metrics, and measurement strategies for assessing cancer care delivery by recognizing the importance of measurement and quality benchmarking in cancer care.^4^ They noted the importance and gap of high-quality “end-of-life” measures and a “significant gap” in measuring what matters most to patients, including patient experiences.^4^ The council also noted persistent issues in implementing and documenting such measures in routine care.^3^

The **conside**RATE questions, a patient-reported measure of serious illness care experience, may address these measurement needs and provide an opportunity to improve patients’ and care partners’ experiences.^5^ This unique measure was created for individuals facing serious illnesses, including those nearing the end of life. It focuses on addressing elements of care that have been identified as meaningful to patients with serious illnesses.^5^ Unlike other measures^6^, the completion time for **conside**RATE is short, ranging from an average of 3 minutes to slightly over 6 minutes, and this measure complements other self-report quality measure, such as The “Heard & Understood” item.^5,7,8^ The **conside**RATE questions is flexible and can be used across settings and diagnoses.^5^ ^9^ Also, the **conside**RATE questions may enhance routine care measurement for cancer patients, provide valuable feedback to care teams, and serve as an outcome measure in research and quality improvement.^5,7^

Preliminary psychometric work on the **conside**RATE questions is promising, but questions remain.^7^ For example, we validated the **conside**RATE questions with older adults (≥ 50 years old) in an online randomized survey using simulated patient experiences.^7^ In this controlled environment, the **conside**RATE questions were reliable and valid. However, the real-world psychometric properties of the **conside**RATE questions are unknown. Importantly, validation with care partners has not yet occurred. Without real-world validation, it remains uncertain whether the **conside**RATE questions accurately capture the serious illness care experience for cancer patients and their care partners.

To address these gaps, we aimed to describe the convergent validity of the **conside**RATE questions when administered to patients and their care partners at a National Cancer Institute (NCI) designated comprehensive cancer center in rural New England.

## Methods

### Design

We conducted a cross-sectional survey study with convenience sampling. We obtained informed consent and provided surveys on iPads or on paper in-person. We reported results using the Checklist for Reporting of Survey Studies (CROSS) (**Appendix 1**).^10^ The Dartmouth Health Institutional Review Board (IRB) approved this study in June 2021 (DH IRB STUDY02000560).

### Participants

We recruited patients with cancer and care partners (family members, caregivers, or friends who accompanied patients) in a National Cancer Institute (NCI) Designated Comprehensive Cancer Center, The Dartmouth Cancer Center (DCC), in Lebanon, New Hampshire.^11^ NCI-Designated Cancer Centers have high survival rates and healthcare quality ratings, making them well-suited for validation.^12,13^ We recruited participants who checked in at the main DCC reception area, where most but not all cancer patients check-in.

To be eligible, participants needed to be a patient or care partner of a patient at the DCC; able to consent and read English; and 18 years of age or older. Parents or guardians of children with cancer were eligible to participate as care partners. We excluded those who could not provide informed consent, those under the age of 18, and persons who were incarcerated.

### Survey validation

We created an online survey using the Qualtrics survey platform.^14^ We designed the survey and analytic plan based on previously conducted online validation studies.^7,15,16^ The 10-minute survey (**Appendix 2**) consisted of 43 questions and included basic demographics, a single item assessing health literacy^17^, and healthcare experience-related questions, including the **conside**RATE questions, and the CANHELP Lite Patient questionnaire.^5,6^

### Survey elements

#### Participant Characteristics

Participants provided demographics, including age, gender, family income, cancer diagnosis, highest level of education, and health literacy.

#### The ConsideRATE Questions

We previously published the development and online validation of **conside**RATE.^7^ Briefly, **conside**RATE is an 8-item, patient-reported experience measure (PREM) that evaluates serious illness care experiences.^5,7^ The **conside**RATE questions can be administered to people in diverse settings, including outpatient, inpatient, and home care. The first seven items are scored on a four-point Likert-like scale ranging from 1 (very bad) to 4 (very good) with a “doesn’t apply” option. The eighth item is an open-ended question that asks, “Are there any other things you want to share?” We did not include the optional ninth question about desired anonymity as it was not relevant for research purposes.

#### CANHELP Lite

CANHELP Lite (Canadian Health Care Evaluation Project) is a 21-item patient-facing and validated serious illness experience measure.^6^ CANHELP Lite consists of items concerning the management of physical and emotional symptoms and the environment of care. All items are scored on a five-point Likert-like scale ranging from 1 (not at all) to 5 (completely) with a “doesn’t apply” option.

### Procedures

We collected data between May 2022 and May 2023. Patients and care partners were approached by research assistants in the waiting room before their scheduled visits, and invited to participate in the study. After informed consent and before their visit, participants completed surveys (iPad, Qualtrics QR code, paper).

### Statistical analyses

Prior to analysis, we removed anyone who did not progress past the first phase of the survey, which included demographics (n=14). Participants did not need to complete all questions on **conside**RATE and CANHELP Lite measures to be included in analyses. We used chi-square analyses to test for mean differences between patients and care partners for selected covariates. In the second analysis phase, we performed psychometric tests of the reliability, discriminant validity, and convergent validity, using a Cronbach’s alpha coefficient for internal consistency and Pearson correlation test, of the **conside**RATE questions across scoring methods. Given the conceptual differences between **conside**RATE and CANHELP Lite and the real-world nature of this study, we interpreted convergent validity statistics, recognizing that perfect alignment was unlikely due to their differing characteristics and the variability in real-world data based on population of interest.^18^ We defined weak correlation as less than r=0.3, moderate between r=0.3 to r=0.7, and strong correlation as at least r=0.7.^19^ We used SPSS Statistics Version 28.0.^20^

#### Overall Instrument Scoring

We tested the convergent validity of the **conside**RATE questions across two scoring approaches: continuous scoring and top-box scoring.^15,16^ For continuous scoring, we used the original **conside**RATE four-point and CANHELP five-point scales. We calculated mean **conside**RATE question scores across all seven items and mean CANHELP Lite scores across all 21 items.^6^ For participants who did not score all questions on either measure, the total number of questions used to calculate the mean was adjusted to equal the total number of answered questions.

For top-box scoring, we dichotomized overall **conside**RATE scores by allocating a “1” to participants who responded “very good” to all measure items they completed, and “0” to participants who did not. We used the original continuous CANHELP scoring. We compared the two measures and tested convergent validity using a point-biserial correlation test.

In addition to continuous and top-box scoring, we initially planned to report on a third method of scoring **conside**RATE, a high-low dichotomization scoring (overall mean scores below the midpoint of 2.5 were “low” and scores 2.5 or higher were “high”). We did not have sufficient participants in the “low” group to reliably report the results of these analyses.

#### Item-by-item scoring: Convergent Validity

To perform item-by-item convergent validity tests of the **conside**RATE questions, two independent reviewers (JN and CHS) matched the seven **conside**RATE questions with seven out of 21 CANHELP Lite questions based on concept. The reviewers resolved discrepancies after review. For continuous scoring, we compared individual items using the **conside**RATE four-point scale to matched items on the CANHELP Lite five-point scale, using Pearson’s correlation tests. For top-box scoring, we compared individual items of **conside**RATE scored using a top-box approach (“very good” vs all else) to match items on the CANHELP Lite five-point scale, using point-biserial correlation tests.

#### Discriminant Validity

To test for discriminant validity, we identified a measure that is conceptually distinct from the serious illness experience measured in **conside**RATE, the validated Single-item Health Literacy (SILs) measure.^17,21,22^ This instrument asks “How confident are you in filling out medical forms by yourself?” and offers the following response options: “Extremely”, “Quite a bit”, “Somewhat”, “A little bit”, “Not at all.” We selected SILs because it does not directly address **conside**RATE constructs or and does overlap conceptually with any of the seven **conside**RATE questions.

#### Subgroup analyses

Overall and item-level convergent validity analyses were conducted across all participants, as well as patient-only and care partner-only subgroups. We also conducted subgroup analyses for participants with low income and participants with low educational attainment. These analyses were based on the overall instrument means. We defined participants with low income as those who earn less than $35,000 annually, for a family of four; this is less than the 138% Federal Poverty Level qualification for Medicaid.^23^ We defined participants with low educational attainment as those who have a high school diploma or equivalent.

### Evaluating outpatient cancer patient experiences

#### Cancer categories

We evaluated patient experience using the **conside**RATE questions continuous scoring approach. First, we selected the two most common cancer diagnoses in our data set and conducted an independent t-test to determine whether there is significant difference in patient experience based on overall **conside**RATE mean score between these two subgroups. Second, we dichotomized all 12 cancer diagnoses associated with reason for visit into “solid” cancer category (e.g., breast cancer, sarcoma) and “hematologic malignancy” cancer category (e.g., leukemia, lymphoma). We conducted an independent t-test to determine whether there is a significant difference in patient experience between these two categories.

#### ConsideRATE open-ended question

Two raters (JN and CHS) reviewed open-text responses to **conside**RATE question eight, “Are there any other things you want to share? Write them here.” We scored responses as positive (1), negative (2), or unclear (3). When comments were mixed, we marked them as unclear. We subsequently compared overall **conside**RATE scores with binary favorability ratings (positive vs negative) by calculating a Phi coefficient for top-box and Pearson coefficient for continuous scores. JN and CHS resolved conflicts through discussion.

## Results

### Participant characteristics

A total of 258 patients and care partners participated in the survey. However, after screening out incomplete surveys, we had a final sample of 244 participants. Approximately half of our participants were patients (n=114, 47%) and the other half were family members or care partners (n=128, 53%). Two participants did not specify patient or care partner roles. Most participants were female (n=170, 70%); one was non-binary. Most participants identified as white or caucasian (n=232, 95%) and non-Hispanic (n=238, 98%). The most common reason for visiting the cancer center was related to blood or marrow transplant (20%) (**Table 1**). Approximately 20% of participants (n=36) chose “doesn’t apply” for one of the seven **conside**RATE questions and 7% of participants (n=36) chose “doesn’t apply” for at least half of the **conside**RATE questions (n=16). Three percent of participants (n=8) chose to skip one of the **conside**RATE questions and no participant chose to skip more than two **conside**RATE questions. Six percent of participants (n = 14) chose to skip at least half of the CANHELP questions.

**Table 1.** Baseline characteristics among patients with cancer and care partner or family member ^1^

|  | Care partner<br>*128, No. (%)* | Patient<br>*114, No. (%)* | P-value<sup>2</sup> |
| --- | --- | --- | --- |
| **Age** |  |  |  |
| 18-34 | 10 (7) | 7 (6.1) | <0.001 |
| 35-54 | 33 (26) | 32 (28) |  |
| 55-74 | 69 (54) | 62 (54) |  |
| 75+ | 16 (13) | 13 (11) |  |
| **Palliative care** |  |  |  |
| No | 81 (63) | 64 (56) | 0.19 |
| Yes | 30 (23) | 25 (22) |  |
| **Gender** |  |  |  |
| Female | 93 (73) | 76 (67) | <0.001 |
| Male | 34 (27) | 38 (33) |  |
| Other | 1 (1) | None |  |
| **Race** |  |  |  |
| American Indian | 3 (2) | 2 (2) | <0.001 |
| Asian | 1 (1) | None |  |
| Black | 1 (1) | None |  |
| White | 121 (95) | 110 (97) |  |
| Other | 1 (1) | 1 (1) |  |
| **Ethnicity** |  |  |  |
| Non-hispanic | 126 (99) | 111 (98) | <0.001 |
| Hispanic | 1 (1) | 2 (2) |  |
| **Cancer diagnoses** <sup>3</sup> |  |  |  |
| Blood or bone marrow | 25 (20) | 24 (21) | 0.76 |
| Breast | 15 (12) | 20 (18) |  |
| Endocrine | 2 (2) | 6 (5) |  |
| Gastroenterology | 14 (11) | 6 (5) |  |
| Genitourinary | 2 (2) | 2 (2) |  |
| Gynecology | 6 (5) | 5 (4) |  |
| Head or Neck | 4 (3) | 3 (3) |  |
| Melanoma | 4 (3) | 5 (4) |  |
| Neuro | 1 (1) | 5 (4) |  |
| Pediatric | 1 (1) | None |  |
| Sarcoma | 2 (2) | 2 (2) |  |
| Thoracic | 5 (4) | 6 (5) |  |
| Unknown | 36 (28) | 25 (23) |  |
| **Health literacy\*** |  |  |  |
| Confident | 115 (90) | 102 (89) | <0.001*** |
| Not confident | 13 (10) | 12 (11) |  |
| **Educational attainment** |  |  |  |
| Some high school or high school/GED degree | 61 (48) | 57 (50) | <0.001 |
| 2-year degree | 16 (13) | 6 (5) |  |
| 4-year degree | 37 (29) | 23 (20) |  |
| Graduate degree** | 14 (11) | 28 (25) |  |
| **Income** |  |  |  |
| \$75,000+ | 51 (40) | 44 (39) | |
| \$50,000 - \$74,999 | 28 (22) | 17 (15) | 0.075 |
| \$25,000 - \$49,999 | 29 (23) | 35 (31) | |
| Less than \$25,000 | 10 (8) | 13 (11) | |
| **Insurance** |  |  |  |
| No | 2 (2) | 1 (1) | <0.001 |
| Yes | 126 (98) | 113 (99) |  |
| **Type of insurance** |  |  |  |
| Employer | 51 (40) | 41 (36) | <0.001 |
| Medicaid | 10 (8) | 14 (12) |  |
| Medicare | 24 (19) | 14 (12) |  |
| Medicare plus supplemental | 27 (21) | 32 (28) |  |
| Purchased | 8 (6) | 6 (5) |  |
| Other | 7 (6) | 7 (6) |  |

<sup>1</sup> Two participants did not specify whether they were care partners or patients
<sup>2</sup> Chi-squared test was performed for categorical variables
<sup>3</sup> We combined (1) “blood or marrow transplant”, (2) “white blood cell cancer”, (3) “blood or bone marrow cancer” into one category called “blood or bone marrow”
\* We converted a 5-item scale (extremely, quite a bit, somewhat, a little bit, not at all) into a 2-item scale (confident or not confident).
\*\* Some college, some high school and high school diplomat were combined into one category
\*\*\* Graduate degree referred to Master’s degree and Doctorate’s degree

### Overall measure

Overall scores using the **conside**RATE questions continuous scoring approach skewed high, with a mean of 3.72 (possible range 1-4) and standard deviation of 0.38 (**Figure 1**). Using a top-box approach, 45% of participants received a top-box score of 1 (indicating the highest possible experience of care rating) and 55% received a top-box score of 0 (less than highest possible rating).

**Figure 1.**
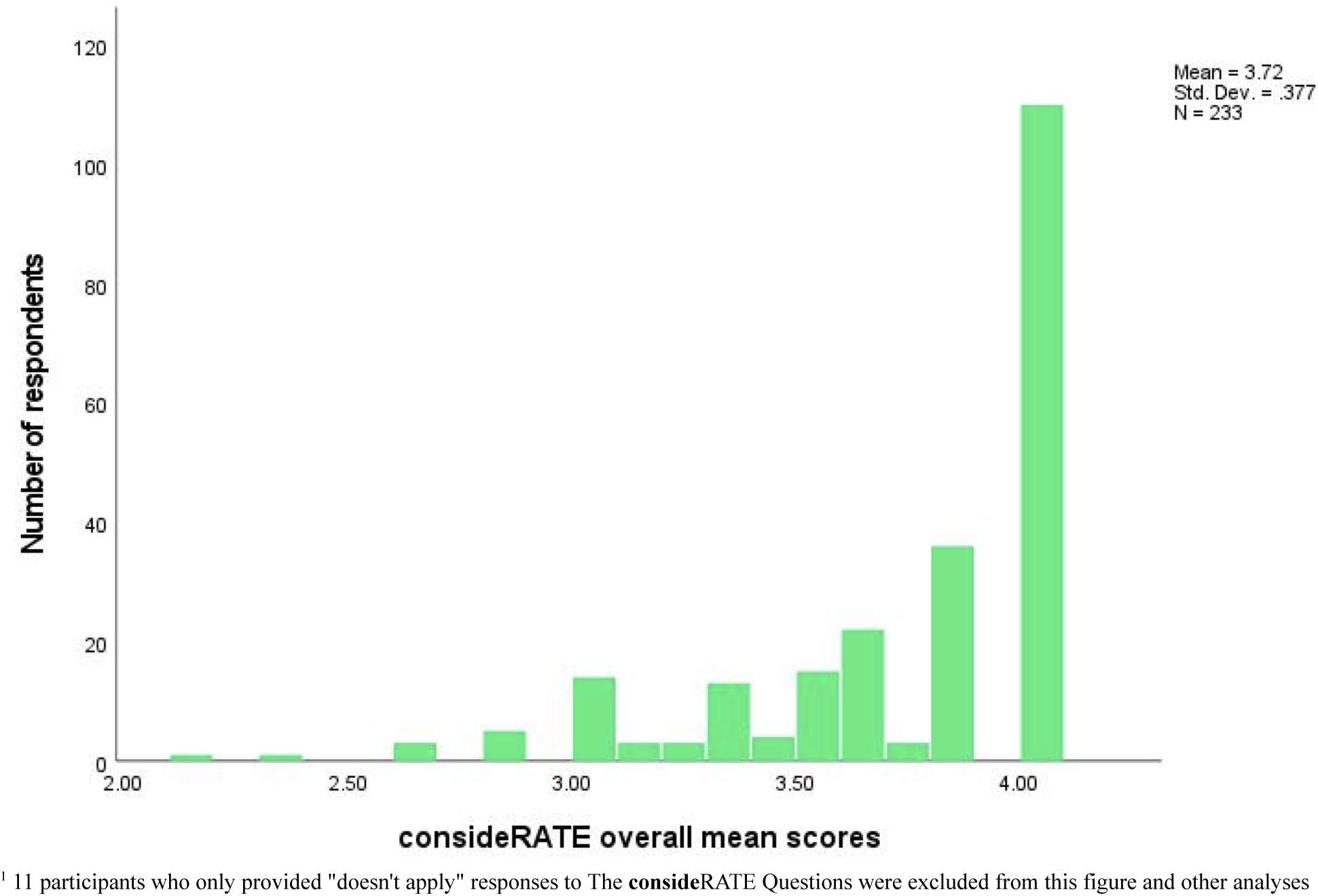
Histogram of **conside**RATE overall mean score per participant (n=233)^1^

### Psychometric assessment

#### Reliability

The **conside**RATE questions demonstrated reliability with internal consistency of 0.86 (**Table 2**). Cronbach’s alpha coefficient for the 7-item measure did not significantly change when an independent item was deleted (**Table 2**).

**Table 2.** Psychometric tests using overall instrument scoring^1^

| Convergent validity | All participants<br>r (p-value) | Patients only<br>r (p-value) | Care partners only<br>r (p-value) | Low-income<br>r (p-value) | Low educational attainment<br>r (p-value) |
| --- | --- | --- | --- | --- | --- |
| Mean **consideRATE** questions scores vs. mean CANHELP Lite score | 0.5 (<0.001)*** | 0.5 (<0.001)*** | 0.5 (<0.001)*** | 0.7 (<0.001)*** | 0.5 (<0.001)*** |
| Top-box dichotomous **consideRATE** questions scores vs. mean CANHELP Lite score | 0.4 (<0.001)*** | 0.5 (<0.001)*** | 0.3 (0.003)** | 0.6 (<0.001)*** | 0.4 (0.005)** |
| Reliability | Mean (SD) | Scale Variance if Item Deleted | Corrected item-Total correlation | Squared Multiple Correlation | Cronbach's Alpha if Item Deleted |
| **Overall Cronbach alpha = 0.864** |  |  |  |  |  |
| Physical item | 3.79 (0.48) | 4.327 | 0.584 | 0.393 | 0.852 |
| Surrounding item | 3.70 (0.49) | 4.346 | 0.558 | 0.329 | 0.856 |
| Feelings item | 3.80 (0.45) | 4.289 | 0.664 | 0.527 | 0.841 |
| What matters most item | 3.84 (0.39) | 4.399 | 0.718 | 0.569 | 0.837 |
| Plan item | 3.78 (0.45) | 4.277 | 0.667 | 0.472 | 0.840 |
| Affairs item | 3.66 (0.50) | 4.174 | 0.630 | 0.476 | 0.846 |
| Expect item | 3.71 (0.47) | 4.234 | 0.655 | 0.452 | 0.842 |

<sup>1</sup>To compute correlation coefficients we used Pearson correlations for continuous scoring and point-biserial correlation tests for top-box scoring

#### Convergent validity: Overall instrument scoring

Among all participants, we found a statistically significant moderate correlation (r=0.5; *p*<0.001) between the overall continuous scores of the **conside**RATE questions and CANHELP Lite, indicating adequate convergent validity. For top-box scoring among all participants, we found a statistically significant weak-moderate correlation (r^pb^=0.4; *p*<0.001) between the overall top-box scoring of the **conside**RATE questions and the CANHELP Lite continuous scores, indicating adequate convergent validity.

For continuous scoring participant subgroup analyses, we found significant correlations ranging from moderate to moderate-high between **conside**RATE and CANHELP Lite: patients only (r=0.5, *p*<0.001); care partners only (r=0.4, *p*<0.001); participants with low income (r=0.7; *p*<0.001); and participants with low educational attainment (r=0.5; *p*<0.001) (**Table 2**). For top-box scoring subgroup analyses, we found significant correlations ranging from weak to moderate: patients only (r^pb^=0.5, *p*<0.001); care partners only (r^pb^=0.3, p=0.003); participants with low income (r^pb^=0.6, *p*<0.001); and participants with low educational attainment (r^pb^=0.4, p=0.005).

#### Convergent validity: Item-by-item scoring

For continuous scoring among all participants, we found statistically significant moderate correlations (ranging from r=0.4 (*p*<0.001) to 0.6 (*p*<0.001)) across all individual matched **conside**RATE continuous scoring and CANHELP Lite questions, indicating adequate convergent validity (**Table 3**). For top-box scoring among all participants, we found statistically significant weak to moderate correlations (ranging from r^pb^=0.3 (*p*<0.001) to r^pb^=0.5 (*p*<0.001)) across all individual questions, indicating adequate convergent validity for most items (**Table 3**).

**Table 3.** Matching **conside**RATE questions with CANHELP Lite questions for convergent and discriminant validity ^1,2^

| Convergent Validity |  |  | ConsiderATE <sup>4</sup> |  |
| --- | --- | --- | --- | --- |
| Construct Summary<br>Care of | ConsiderATE questions | CANHELP Lite questions | Continuous<br>r (p-value) | Top-box<br>r (p-value) |
| Physical problems | Q1: "How would you rate our attention to your physical problems? Things like pain, dry mouth or trouble breathing" | Q1: "How satisfied are you that physical symptoms you had (for example: pain, shortness of breath, nausea) were adequately assessed and controlled?" | 0.5 (<0.001)*** | 0.4 (<0.001)*** |
| Emotional problems | Q2: "How would you rate our attention to your feelings? Things like feeling sad, worried, or like a burden" | Q2: "How satisfied are you that emotional problems you had (for example: depression, anxiety) were adequately assessed and controlled?" | 0.5 (<0.001)*** | 0.4 (<0.001)*** |
| Surrounding problems | Q3: "How would you rate our attention to your surroundings? Things like noise, light, or warmth" | Q3: "How satisfied are you with the environment or the surroundings in which you were cared for?" | 0.6 (<0.001)*** | 0.5 (<0.001)*** |
| What matters most | Q4: "How would you rate our respect for what matters to you? Things like values, preferences about care, or important activities" | Q4: "How satisfied are you that you received consistent information about your condition from all doctors and nurses looking after you?" | 0.6 (<0.001)*** | 0.4 (<0.001)*** |
| Plans | Q5: "How would you rate our communication about your plans? Things like medicines, procedures or place of care" | Q5: "How satisfied are you with discussions with your doctor(s) about where you would be cared for (in hospital, at home, or elsewhere) if your condition worsened?" | 0.5 (<0.001)*** | 0.4 (<0.001)*** |
| Affairs | Q6: "How would you rate our attention to your affairs? Things like a will, finances, or advance directives for care" | Q6: "How satisfied are you that you were able to manage the financial costs associated with your illness?" | 0.4 (<0.001)*** | 0.3 (<0.001)*** |
| What to expect | Q7: "How would you rate our communication about what you can expect? Things like illness getting worse or time left to live" | Q7: "How satisfied are you with discussions with your doctor(s) about the use of life-sustaining technologies (for example: CPR or cardiopulmonary resuscitation, breathing machines, dialysis)?" | 0.6 (<0.001)*** | 0.5 (<0.001)*** |
| **Discriminant Validity** | **ConsiderATE questions** | **Single-item Health Literacy <sup>3</sup>** |  |  |
| Physical problems | Q1: "How would you rate our attention to your physical problems? Things like pain, dry mouth or trouble breathing" | Q1: "How confident are you in filling out medical forms by yourself?" | -0.02 (0.742) | 0.004 (0.948) |
| Emotional problems | Q2: "How would you rate our attention to your feelings? Things like feeling sad, worried, or like a burden" | Q1: "How confident are you in filling out medical forms by yourself?" | -0.01 (0.873) | 0.006 (0.925) |
| Surrounding problems | Q3: "How would you rate our attention to your surroundings? Things like noise, light, or warmth" | Q1: "How confident are you in filling out medical forms by yourself?" | 0.07 (0.280) | 0.03 (0.682) |
| What matters most | Q4: "How would you rate our respect for what matters to you? Things like values, preferences about care, or important activities" | Q1: "How confident are you in filling out medical forms by yourself?" | -0.05 (0.504) | -0.02 (0.711) |
| Plans | Q5: "How would you rate our communication about your plans? Things like medicines, procedures or place of care" | Q1: "How confident are you in filling out medical forms by yourself?" | 0.09 (0.198) | 0.09 (0.184) |
| Affairs | Q6: "How would you rate our attention to your affairs? Things like a will, finances, or advance directives for care" | Q1: "How confident are you in filling out medical forms by yourself?" | 0.04 (0.564) | 0.004 (0.956) |
| What to expect | Q7: "How would you rate our communication about what you can expect? Things like illness getting worse or time left to live" | Q1: "How confident are you in filling out medical forms by yourself?" | -0.006 (0.933) | 0.02 (0.729) |

<sup>1</sup> To compute correlation coefficients we used Pearson correlation tests for continuous scoring and point-biserial correlation tests for top-box scoring.
<sup>2</sup> All r coefficients were rounded to the nearest tenths
<sup>3</sup> The single-item health literacy measure asks "How confident are you in filling out medical forms by yourself?" and offers the following response options: "Extremely", "Quite a bit", "Somewhat", "A little bit", "Not at all."
<sup>4</sup> We rounded all Pearson's correlation coefficients to the nearest tenth.
\*\*\* p<0.001, \*\* p< 0.01, \* p< 0.05

#### Discriminant validity

We found no significant correlations for **conside**RATE and SIL measure, indicating adequate discriminant validity (**Table 3**). We found no significant correlations across most mismatched items, indicating discriminant validity (**Table 3**).

#### Convergent validity: Subgroup analyses

For continuous scoring, item-level analyses for participant subgroups were mostly statistically significant, with weak to high correlations (**Appendix 3**). We did not find significant correlations between matched items for **conside**RATE Q1 “physical problems” for the patients only subgroup, Q2 “emotional problems” for participants with low educational attainment, and Q4 “what matters most” for participants with low educational attainment (**Appendix 3**). For top-box scoring, item-level analyses for participant subgroups were mostly statistically significant, with weak to moderate correlations (**Appendix 3**). For participants with low educational attainment, we did not find significant correlations between matched items for **conside**RATE Q2 “emotional problems”, Q3 “surrounding problems”, Q4 “what matters most”, and Q6 “affairs” (**Appendix 3**).

### Evaluating patient experience

#### ConsideRATE overall scoring

The two most common reported reasons for visit were for “blood or bone marrow” and “breast cancer” appointments. Participants who reported being present for a “blood or bone marrow” appointment (n = 49) received an overall **conside**RATE mean score of 3.68 and those reporting being present for a “breast cancer” (n = 35) appointment received an overall **conside**RATE mean score of 3.72. We found no significant difference between these two overall mean scores (p = 0.617). We found no significant difference between participants who reported a “solid” cancer category for reason for visit (n = 116, mean score = 3.71) and those who reported a “hematologic malignancy” cancer category (n = 49, mean score = 3.68) (p = 0.62).

#### ConsideRATE open-ended question

181 patients and care partners provided free-text responses. Agreement between the two reviewers was 88% (Weighted kappa=0.972). We received 36 positive comments, 17 negative comments, and 10 unclear comments. The correlation coefficient between free-text comments (positive, negative) and overall mean **conside**RATE scores were not statistically significant for continuous (r=0.2, p=0.236) or top-box (phi=0.1, p=0.328) scoring.

In cases where the **conside**RATE score was positive and the comment was negative, participants often offered specific constructive criticism in areas beyond the scope of the questionnaire (**Appendix 4**). The most negative **conside**RATE scores did not come with associated open-text critiques as we received no comments from participants who had an overall **conside**RATE mean score less than 3 **(Appendix 4)**.

## Discussion

In the first real-world clinical and psychometric test of the **conside**RATE questions, a brief, plain language measure rooted in the constructs of importance to patients and care partners, we found adequate convergent validity in a routine outpatient cancer care setting.

We conducted three psychometric assessments: convergent validity, discriminant validity, and internal reliability. For convergent validity assessment among all participants and all other subgroups, continuously-scored **conside**RATE demonstrated stronger convergent validity compared to top-box **conside**RATE scores (sometimes known as proportion of top scores or top score analysis). However, top-box scores had acceptable validity and may be easier for clinical teams to interpret and use in routine care. While continuous **conside**RATE scores among care partners-only subgroups revealed moderate convergent validity consistent with continuously scored patients-only subgroups, top-box **conside**RATE scores among care partners-only subgroups revealed slight decrease in correlation coefficient for convergent validity inconsistent with top-box scored patient-only subgroups. Although we found care partner scores to be valid broadly, it is interesting that we found a lower convergent correlation between **conside**RATE top-box and CANHELP Lite scores specifically among care partners-only. For discriminant validity assessment, we found no correlation between our health measure **conside**RATE and single-item health literacy measure. For internal reliability assessment, we found high internal consistency in the **conside**RATE questions. This is expected because **conside**RATE is a healthcare experience measure and each health-related item is likely to be positively correlated with the overall health construct being evaluated.^24^

These results provide patients and care partners’ direct assessment of their cancer care experiences, an essential step in instrument validation.^25^ The results of this study are consistent with previous **conside**RATE simulated online validation work and support the continued use of **conside**RATE in routine care and research.^7^ The association between **conside**RATE and CANHELP Lite was less strong than that seen in our previous simulated online validation work, which likely was due to the previous study’s highly controlled nature.^7^ Given the imperfect comparison between the two measures, along with the expected variation within this real-world sample of patients and care partners, these scores may provide sufficient evidence to support convergent validity in these populations.^6,7^ Results of the current study not only contribute a pragmatic assessment of **conside**RATE, but suggest that an alternative top-box method of scoring is also acceptable, particularly when used with patients. Given ceiling effects in continuous **conside**RATE scores, top-box scoring may provide a more useful and meaningful metric for routine assessment of serious illness care experience.^15,25–27^

Our previous work generated evidence of the **conside**RATE questions’ psychometric validity in an online study.^7^ Importantly, the current study marks our first use of the **conside**RATE questions in a clinical setting, specifically at an NCI-Designated cancer center. This study also demonstrates the use of the **conside**RATE questions with a wide age range, from young adults (18 years and older) to older adults (75 years and older).

### Implications (Clinical and Research)

The **conside**RATE questions fill an important gap in the literature for psychosocial care providers. The “Measuring What Matters” project (MWM), a joint endeavor from the American Academy of Hospice and Palliative Medicine and the Hospice and Palliative Nurses Association, identified a need for measures of patient/family care experience in serious illness.^28^ Like the CANHELP and CANHELP Lite, **conside**RATE measures the subjective experiences of care quality but does so with a particular focus on accessibility for those with serious illnesses and potentially associated cognitive impairments, by providing visual icons next to their relevant texts.^5,7^ The **conside**RATE questions may complement the instrument developed by Gramling and colleagues, “Feeling Heard and Understood”.^8^ Concerning cancer care, a move towards “Measuring What Matters,” may be a welcome additive to discrete outcome and experience measures, as well as quality of life measures. Furthermore, the **conside**RATE questions contribute to the mission of the Cancer Outcomes Measurement Working Group (COMWG) and its call to identify future research priorities concerning patient needs and satisfaction.^29^ Given consideRATE measures discrete outcomes that matter most to patients and care partners, but also allows them to offer their subjective interpretation, this measure may be consistent with that call.

The **conside**RATE questions are being studied, adapted and implemented in diverse clinical settings relevant to serious illness populations, including those with an inpatient focus, a specialty palliative care focus, and potentially mixed diagnosis.^30–32^ This increased activity is generating opportunities for future benchmarking and comparisons across various clinical contexts. In addition to feasibility and acceptability studies, **conside**RATE is being used as an outcome in studies of serious illness.^7^ Beyond research applications, and perhaps particularly important for psychosocial care in oncology, **conside**RATE has proven to be both feasible and useful as a service recovery mechanism in routine care by addressing patient concerns and satisfaction, thereby helping to improve the overall care experience.^33^

Patient-reported experience measures (PREMs) can be described as opportunities to describe objective patient experiences and patient satisfaction. There is a distinction between patient experiences and satisfaction that is well-described by Bull and colleagues.^34–36^ The utility of **conside**RATE may be at least in part because, unlike many similar measures, it is not strictly a measure of patient experience or satisfaction, but a marriage of the two, centering on the psychological and social needs of patients (**Figure 2**).^32,37–39^ Although these types of blended experience and satisfaction measures may exist, there is little work to describe these measures as conceptually different from satisfaction or experience measures. We believe that to rigorously measure and improve the complex care of patients, particularly those with serious illness, there must be a multiplicity of measures and outcomes to allow researchers and clinicians to effectively measure and improve what matters most.

**Figure 2.**
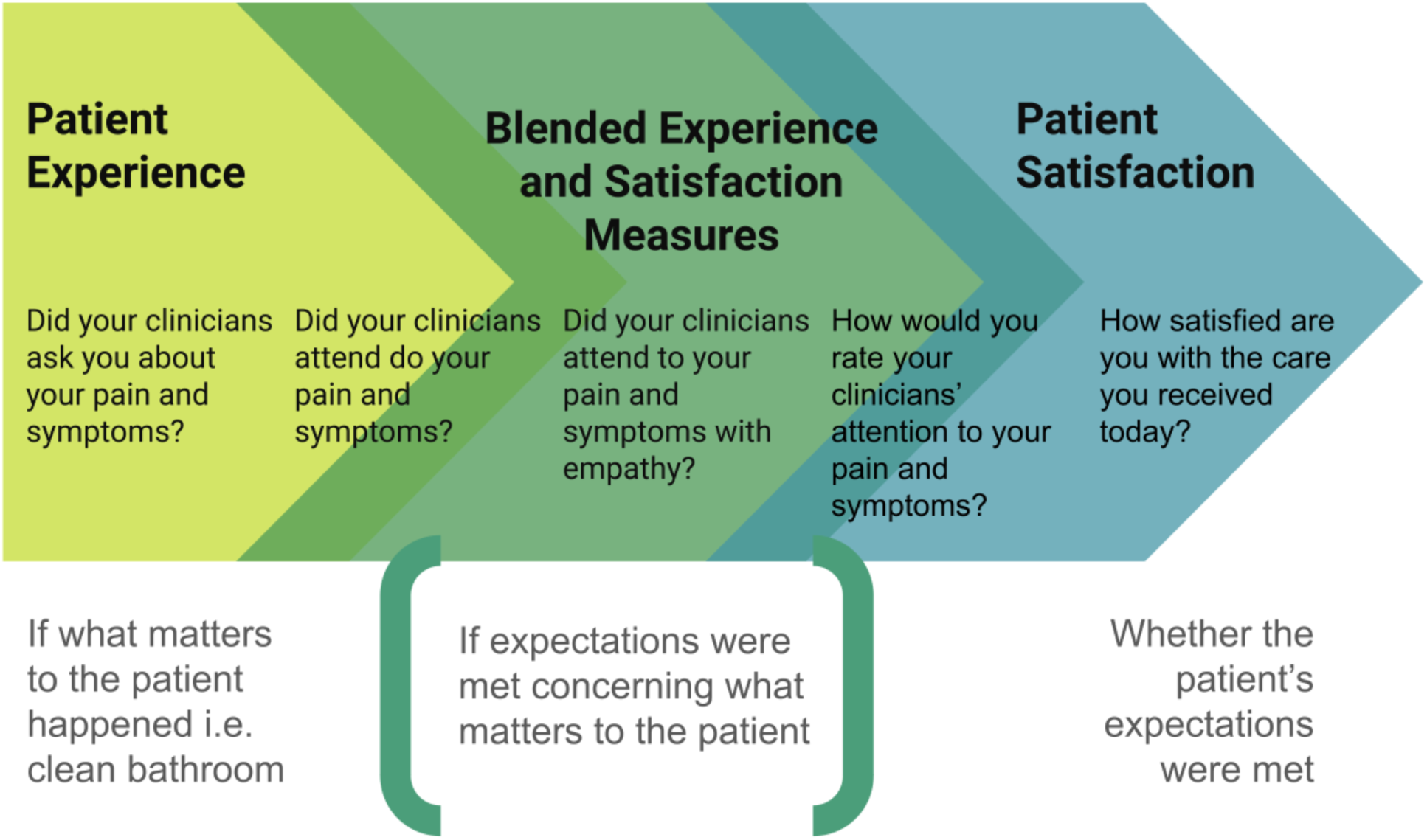
The experience and satisfaction measure spectrum

### Limitations

Recruiting participants in-person may have led to a social desirability bias among participants and unconscious selection bias by the recruitment team. COVID-19 may have influenced recruitment due to universal masking and increased telehealth visits. Additionally, administering the CANHELP Lite, a much longer instrument, in addition to **conside**RATE may have been burdensome to patients and led to rushed completion or incomplete surveys.^5,6^ To validate the individual items of **conside**RATE within the meta-construct of serious illness care experience, we matched them with individual CANHELP Lite scores.^7^ This is not a standard use of the CANHELP Lite measure; therefore, the item-level validity should be understood within that context. Additionally, we did not exclude **conside**RATE responses for participants that answered at least one item, potentially influencing the **conside**RATE overall individual score for these particular participants by overestimating or underestimating associations depending on the patterns of missing responses. The CANHELP Lite developers recommend excluding missing data in cases where more than half of responses are missing, but for congruity we did not adopt this approach in our study. We were unable to determine how similar patient and care partners dyad scores were on **conside**RATE due to the anonymity of the survey. The generalizability of our results may be limited by the inclusion and exclusion criteria used in this study and the fact that data were collected from a single site. Future studies in multiple types of settings with diverse populations will be needed. Also, future studies may definitely determine whether care partners might be able to function as proxy measure completers in some situations.

## Conclusion

We think there is sufficient evidence to support the use of **conside**RATE by both psychosocial researchers and clinicians working in the area of serious illness, including with oncology populations. In this context, **conside**RATE could be used to measure the effectiveness of research and quality improvement interventions, as well as to monitor care quality across clinical contexts and serve as a mechanism for service recovery.

## CRediT authorship contribution statement

**Joseph P. Nano**: recruitment, methodology, formal analysis, and writing the original draft. **Gabrielle Stevens**: methodology, formal analysis, writing – review and editing. **Glyn Elwyn:** conceptualization, writing – review and editing. **Leeza Petrov:** recruitment, writing – review and editing. **Saahithya Gowrishankar:** recruitment, writing – review and editing. **Shaday Robitaille**: recruitment, writing – review and editing. **Arrica D. Van Citters**: conceptualization, writing – review and editing. **Eugene Nelson:** conceptualization, writing – review and editing. **Garrett T. Wasp:** writing – review and editing. **Kathryn Kirkland:** conceptualization, writing – review and editing. **Meredith A. MacMartin:** writing – review and editing. **Catherine H. Saunders:** conceptualization, methodology, formal analysis, investigation, draft – review and editing.

## Data availability statement

The data which supports the findings of this research are available upon request of the authors.

### Acknowledgments

We are grateful to the patients and care partners who participated in this study. We also thank the Dartmouth Cancer Center healthcare staff and receptionists for their cooperation and support during recruitment. We are tremendously grateful to our research coordinator, Cheryl A. Page, without whom this work would have been very difficult. We also extend our thanks to select members of the Healthcare Experience (HEx Lab) research group for providing feedback on our manuscript: Ailyn Sierpe, Anji Zhu, Boyoung Ahn, Emily Zhang, and Rhea Sachdeva.

